# A Population-Based Study of Plasma Metabolomic Profiles of Persistent Tinnitus Identifies Candidate Biomarkers

**DOI:** 10.1101/2022.06.11.22276279

**Authors:** Oana A. Zeleznik, D. Bradley Welling, Konstantina Stankovic, Lisa Frueh, Raji Balasubramanian, Gary C. Curhan, Sharon G. Curhan

## Abstract

Tinnitus, the perception of sound without the corresponding external stimulus, currently affects 15% of the US population. There is a critical need to unravel the heterogeneous etiologies underlying tinnitus and identify tailored treatment targets. Metabolomic studies have elucidated biological pathways underlying several neurodegenerative disorders and could identify metabolic perturbations that influence tinnitus development. We conducted the first large-scale metabolomic study to identify novel tinnitus biomarkers. We cross-sectionally evaluated plasma metabolite profiles and persistent tinnitus among 6477 women (488 with daily tinnitus ≥5 minutes and 5989 controls). A broad array of 466 metabolites was measured using liquid-chromatography mass spectrometry. Logistic regression was used to estimate odds ratios (OR, per 1 SD increase in metabolite values) and 95% confidence intervals (CI) for each individual metabolite while Metabolite Set Enrichment Analysis (MSEA) was used to identify metabolite classes enriched for associations with tinnitus; all models were adjusted for multiple comparisons. Compared with controls, homocitrulline (OR(95%CI)=1.32(1.16-1.50), C38:6 phosphatidylethanolamine (PE; 1.24(1.12-1.38)), C52:6 triglyceride (TAG; 1.22(1.10-1.36)), C36:4 PE (1.22(1.1-1.35)), C40:6 PE (1.22(1.09-1.35)), and C56:7 TAG (1.21(1.09-1.34)) were positively associated, whereas alpha-keto-beta-methylvalerate (0.68(0.56-0.82)) and levulinate (0.60(0.46-0.79)) were inversely associated with tinnitus (adjusted-p<0.05). Among metabolite classes, TAGs, PEs, and diglycerides (DAGs) were positively associated, while phosphatidylcholine (PC) plasmalogens, lysophosphatidylcholines (LPC), and cholesteryl esters were inversely associated with tinnitus (false discovery rate <0.05). This study identified novel plasma metabolites and metabolite classes that were significantly associated with persistent tinnitus. These findings extend our current understanding of tinnitus and could inform investigations of therapeutic targets for this challenging disorder.

## Introduction

Tinnitus is the perception of sound in the absence of an external stimulus. ^1^ Persistent tinnitus can be disabling, adversely impacting sleep, work, daily function, and quality of life. ^2^ In the US, approximately 50 million individuals suffer from tinnitus, among whom 3 million are severely disabled by it. ^3^ The etiology of tinnitus is often unknown, and for most individuals the effectiveness of proposed treatments is uncertain. ^4^ Therefore, identifying potentially modifiable risk factors could aid in prevention and inform targeted treatments for this challenging condition.

There is a critical need to improve the understanding of the biologic underpinnings of tinnitus and the diverse underlying etiologies. ^5^ Accumulating evidence suggests complex interactions between individual-level and environmental factors influence the generation and persistence of tinnitus. ^6^ Metabolomic studies provide a comprehensive picture of an individual’s metabolic status and have elucidated some of the biological pathways underlying several neurodegenerative conditions. Thus, metabolomic assays are powerful tools to identify disease biomarkers and pathoetiologic processes. The metabolome encompasses the collection of small molecules in biologic samples, including organic acids, amino acids, nucleotides, sugars, lipids, and acylcarnitines. Metabolite levels are ‘downstream’ of transcriptional and translational processes and reflect direct input from the diet, environment and intestinal microbiome, thus the systematic analysis of metabolites can uncover valuable insights into a condition as complex as tinnitus.

Previous studies have identified metabolomic biomarkers for neurodegenerative conditions such as Parkinson’s disease, Alzheimer’s and other dementias, and Huntington’s disease, ^7,8^ but published metabolomic studies of persistent tinnitus in humans are lacking. A study in rats examined plasma metabolic profiles associated with acoustic trauma, hyperacusis, and tinnitus and identified several metabolic pathways, including the urea cycle, tricarboxylic acid (TCA) cycle, methionine cycle and purine and pyrimidine metabolism. ^9^ Notably, a number of metabolite alterations demonstrated in brain tissue samples were also demonstrated in plasma. ^9^ However, no human studies have evaluated plasma metabolomic profiles and persistent tinnitus. Therefore, we conducted a large, cross-sectional study in two well-characterized cohorts of US women, the Nurses’ Health Study (NHS) and Nurses’ Health Study II (NHSII), to identify metabolomic alterations associated with prevalent persistent tinnitus.

## Methods

### Study Populations

In 1976, 121,700 female registered nurses aged 30-55 years enrolled in the NHS with the return of a mailed questionnaire. ^10^ Participants have been followed biennially with questionnaires on reproductive history, lifestyle factors, diet, medication use, and new disease diagnoses. In 1989–1990, 32,826 NHS participants provided blood samples and completed a short questionnaire. Briefly, women arranged to have their blood drawn (two 15 mL sodium heparin tubes) and shipped with an ice pack, via overnight courier, to our laboratory, where it was processed and separated into plasma, red blood cell, and white blood cell components and frozen in gasketed cryovials in the vapor phase of liquid nitrogen freezers. Using the same protocol, 18,743 women provided a second blood sample between 2000-2002.

The NHSII began in 1989 with 116,429 female registered nurses aged 25-42 years, with biennial follow-up using similar questionnaires as NHS. Between 1996 and 1999, 29,611 NHSII participants provided blood samples and completed a short questionnaire. ^11^ Premenopausal women (*n*=18,521) who had not taken hormones, been pregnant, or lactated within the past 6 months provided blood samples drawn 7–9 days before the anticipated start of their next menstrual cycle (luteal phase). All other women (e.g., premenopausal taking hormones or postmenopausal; *n*=11,090) provided a single 30 mL untimed blood sample. Between 2010-2012, 17,275 women provided a second blood sample. Samples were shipped and processed identically to the NHS samples.

The study protocol was approved by the institutional review boards of the Brigham and Women’s Hospital and Harvard T.H. Chan School of Public Health. The return of the self-administered questionnaire and blood sample was considered to imply consent.

### Tinnitus assessment

Information on tinnitus was collected on the biennial questionnaires in 2012 and 2016 in NHS, in 2009, 2013 and 2017 in NHS II, and in 2010-2012 on the hearing study supplemental questionnaires (HSSQ) in subcohorts of NHS and NHSII. These questionnaires included the question: “In the past 12 months, have you had ringing, roaring, or buzzing in your ears or head?” The six response options ranged from never to daily. Information on age of onset, how long the sounds last, and whether the sound affects the participant’s ability to sleep, work, concentrate, or engage in daily activities was also collected. The outcome in this study was self-reported persistent daily tinnitus; a case was defined as tinnitus experienced daily and lasting ≥5 minutes for one year or longer (for brevity, hereafter referred to as ‘tinnitus’). We examined a stringent definition of tinnitus to reduce potential misclassification and focus on cases of tinnitus that were most likely to be clinically meaningful. A control was defined as “never” tinnitus. This study included 488 prevalent tinnitus cases and 5989 controls. For all cases, the metabolomic measurements were obtained from blood samples collected after the date of the participant’s first report of persistent tinnitus.

### Assessment of covariates

Information on covariates was obtained from substudy questionnaires completed at the time of blood collection and from biennial questionnaires completed nearest to and before the date of the blood collection. We adjusted for cohort, collection, endpoints from the previously performed metabolomics studies (Supplementary Table 1; see next paragraph on Metabolite profiling), age (continuous?), fasting status, season, menopausal status, hormone use and oral contraceptive use at time of blood draw, as well as factors potentially associated with risk of tinnitus, using updated information on body mass index (continuous), physical activity (metabolic equivalents of task-hours/week, continuous), smoking, hypertension, diabetes mellitus, dietary intake (the Dietary Approaches to Hypertension (DASH) adherence score, a validated measure of adherence to a healthy dietary pattern demonstrated to lower blood pressure and associated with lower risk of hypertension, cardiovascular disease, diabetes, and cognitive decline), ^12^ alcohol intake, depression (clinician-diagnosed or use of antidepressant medications), and self-reported hearing status. Covariate information on many of these factors has been formally validated. ^13-17^

### Metabolite profiling

Metabolomics assays were performed within 15 prospective case-controls studies nested within the NHS and NHSII, including different cancers (breast, ovarian, colon, pancreatic), Diabetes, Stroke, Glaucoma, Inflammatory bowel disease, Amyotrophic lateral sclerosis, Parkinson’s disease, Rheumatoid arthritis, post-traumatic stress disorder. Supplementary Table 1 shows the number of tinnitus cases and controls by nested case-control study. We used the probit transformation within individual studies to correct for batch effects before merging individual metabolomic data across studies. In total, 6477 women responded to the tinnitus questions and had metabolomic data previously measured. Plasma metabolites were profiled at the Broad Institute of MIT and Harvard (Cambridge, MA) using three complimentary liquid chromatography tandem mass spectrometry (LC-MS/MS) methods designed to measure amino acids, amines, lipids, free fatty acids, bile acids, sugars, organic acids, purines, and pyrimidines, acids as described previously. ^18-22^ For each method, pooled plasma reference samples were included every 20 samples and results were standardized using the ratio of the value of the sample to the value of the nearest pooled reference multiplied by the median of all reference values for the metabolite. Within a study, samples from the two cohorts were run together, with matched case-control sets distributed randomly within the batch, and the order of the case and controls within each set randomly assigned. Therefore, the case and its control were always directly adjacent to each other in the analytic run, thereby limiting variability in platform performance across matched case-control pairs. In addition, 2257 quality control (QC) samples, to which the laboratory was blinded, were also profiled. These were randomly distributed among the participants’ samples.

Hydrophilic interaction liquid chromatography (HILIC) analyses of water soluble metabolites in the positive ionization mode were conducted using an LC-MS system comprised of a Shimadzu Nexera X2 U-HPLC (Shimadzu Corp.; Marlborough, MA) coupled to a Q Exactive mass spectrometer (Thermo Fisher Scientific; Waltham, MA). We refer to this method as HILIC-positive. Metabolites were extracted from plasma (10 µL) using 90 µL of acetonitrile/methanol/formic acid (74.9:24.9:0.2 v/v/v) containing stable isotope-labeled internal standards (valine-d8, Sigma-Aldrich; St. Louis, MO; and phenylalanine-d8, Cambridge Isotope Laboratories; Andover, MA). The samples were centrifuged (10 min, 9,000 x g, 4°C), and the supernatants were injected directly onto a 150 × 2 mm, 3 µm Atlantis HILIC column (Waters; Milford, MA). The column was eluted isocratically at a flow rate of 250 µL/min with 5% mobile phase A (10 mM ammonium formate and 0.1% formic acid in water) for 0.5 minute followed by a linear gradient to 40% mobile phase B (acetonitrile with 0.1% formic acid) over 10 minutes. MS analyses were carried out using electrospray ionization in the positive ion mode using full scan analysis over 70-800 m/z at 70,000 resolution and 3 Hz data acquisition rate. Other MS settings were: sheath gas 40, sweep gas 2, spray voltage 3.5 kV, capillary temperature 350°C, S-lens RF 40, heater temperature 300°C, micro scans 1, automatic gain control target 1e6, and maximum ion time 250 ms.

Negative ionization mode data were acquired using an ACQUITY UPLC (Waters) coupled to a 5500 QTRAP triple quadrupole mass spectrometer (AB SCIEX) running a modified version of the hydrophilic interaction chromatography (HILIC) method described by Bajad et al. ^22^ We refer to this method as HILIC-negative. Plasma samples (30 μL) were extracted using 120 μL of 80% methanol (VWR) containing 0.05 ng/μL inosine-15N4, 0.05 ng/μL thymine-d4, and 0.1 ng/μL glycocholate-d4 as internal standards (Cambridge Isotope Laboratories). The samples were centrifuged (10 min, 9000g, 4 °C) and the supernatants (10 μL) were injected directly onto a 150×2.0–mm Luna NH2 column (Phenomenex) that was eluted at a flow rate of 400 μL/min with initial conditions of 10% mobile phase A [20 mmol/L ammonium acetate and 20 mmol/L ammonium hydroxide (Sigma-Aldrich) in water (VWR)] and 90% mobile phase B [10 mmol/L ammonium hydroxide in 75:25 v/v acetonitrile/methanol (VWR)] followed by a 10-min linear gradient to 100% mobile phase A. The ion spray voltage was -4.5 kV and the source temperature was 500 °C.

Plasma lipids were profiled using a Shimadzu Nexera X2 U-HPLC (Shimadzu Corp.; Marlborough, MA). We refer to this method as C8-positive. Lipids were extracted from plasma (10 µL) using 190 µL of isopropanol containing 1,2-didodecanoyl-sn-glycero-3-phosphocholine (Avanti Polar Lipids; Alabaster, AL). After centrifugation, supernatants were injected directly onto a 100 × 2.1 mm, 1.7 µm ACQUITY BEH C8 column (Waters; Milford, MA). The column was eluted isocratically with 80% mobile phase A (95:5:0.1 vol/vol/vol 10mM ammonium acetate/methanol/formic acid) for 1 minute followed by a linear gradient to 80% mobile-phase B (99.9:0.1 vol/vol methanol/formic acid) over 2 minutes, a linear gradient to 100% mobile phase B over 7 minutes, then 3 minutes at 100% mobile-phase B. MS analyses were carried out using electrospray ionization in the positive ion mode using full scan analysis over 200–1100 m/z at 70,000 resolution and 3 Hz data acquisition rate. Other MS settings were: sheath gas 50, in source CID 5 eV, sweep gas 5, spray voltage 3 kV, capillary temperature 300°C, S-lens RF 60, heater temperature 300°C, microscans 1, automatic gain control target 1e6, and maximum ion time 100 ms. Lipid identities were denoted by total acyl carbon number and total double bond number.

Metabolites of intermediate polarity, including free fatty acids and bile acids, were profiled using a Nexera X2 U-HPLC (Shimadzu Corp.; Marlborough, MA) coupled to a Q Exactive (Thermo Fisher Scientific; Waltham, MA). We refer to this method as C18-negative. Plasma samples (30 µL) were extracted using 90 µL of methanol containing PGE2-d4 as an internal standard (Cayman Chemical Co.; Ann Arbor, MI) and centrifuged (10 min, 9,000 x g, 4°C). The supernatants (10 µL) were injected onto a 150 × 2.1 mm ACQUITY BEH C18 column (Waters; Milford, MA). The column was eluted isocratically at a flow rate of 450 µL/min with 20% mobile phase A (0.01% formic acid in water) for 3 minutes followed by a linear gradient to 100% mobile phase B (0.01% acetic acid in acetonitril) over 12 minutes. MS analyses were carried out using electrospray ionization in the negative ion mode using full scan analysis over m/z 70-850. Additional MS settings are: ion spray voltage, -3.5 kV; capillary temperature, 320°C; probe heater temperature, 300 °C; sheath gas, 45; auxiliary gas, 10; and S-lens RF level 60.

Raw data from orbitrap mass spectrometers were processed using TraceFinder 3.3 software (Thermo Fisher Scientific; Waltham, MA) and Progenesis QI (Nonlinear Dynamics; Newcastle upon Tyne, UK) and targeted data from the QTRAP 5500 system were processed using MultiQuant (version 2.1, SCIEX; Framingham, MA). For each method, metabolite identities were confirmed using authentic reference standards or reference samples.

During QC analysis, 192 metabolites were excluded because they were not robust with respect to sample collection characteristics ^21^ while additional 132 metabolites were excluded because they were measured in <100 participants. In total, 466 metabolites were included in the study. Among the 466 analyzed metabolites, 389 (83%) metabolites had a coefficient of variation ≤25%. Metabolite sample sizes varied between n=100 and n=6477 which was mostly due to the selected extraction methods in the individual nested case-control studies (up to 4 methods selected from HILIC-positive, HILIC-negative, C8-positive, C18-negative) and, to a lesser extent, missing metabolite values.

### Statistical analysis

Missing values for metabolites with <25% missingness were imputed with 1/2 of the minimum value measured for that metabolite. Metabolites missing in ≥25% of the samples were evaluated among samples with non-missing values only. Continuous metabolite values were transformed to probit scores within a study to correct for batch effects, reduce the influence of skewed distributions and heavy tails on the results, and to scale the measured metabolite values to the same range. Logistic regression was used to evaluate metabolite associations, modeled continuously, with tinnitus. We present the odds ratios (OR) and 95% confidence intervals (95% CI) for an increase of 1 standard deviation (SD) in probit-transformed metabolite levels estimated with four statistical models: Model 1 adjusts for age, race, fasting status, season of blood draw, collection, cohort, endpoint; Model 2 adjusts for all covariates in Model 1+ body mass index; Model 3 adjusts for all covariates in Model 2 + menopausal status + current oral hormone use, current oral contraceptive use; and Model 4 adjusts for all covariates in Model 3 + smoking status, physical activity, DASH dietary score, alcohol intake. We used the number of effective tests (NEF) to account for testing multiple correlated hypotheses.^23^ We used metabolite set enrichment analysis (MSEA) ^24^ and the false discovery rate (FDR) ^25^ to identify metabolite classes enriched for associations with tinnitus. We present and discuss individual metabolites with NEF-corrected p<0.2 and metabolite classes with FDR<0.2. All analyses were performed with R version 4.0.3. ^26^

The main analysis included 488 cases (defined as daily tinnitus lasting ≥5 minutes) and 5989 controls (no tinnitus). We conducted stratified analyses restricted to those without self-reported hearing loss (129 cases and 3928 controls) and to those with moderate or severe hearing loss (160 cases and 675 controls). In sensitivity analyses, we restricted to women who participated in the first NHS collection (268 cases and 2683 controls), the second NHS collection (63 cases and 1128 controls), the first NHSII collection (147 cases and 2021 controls), and to controls as defined within an individual study (235 cases and 3021 controls; i.e., controls in the breast cancer or ovarian cancer prospective case-control study).

## Results

### Study population

Characteristics of the study population at the time of the blood collection are presented in Table 1. Among those with metabolomic profiles who responded to the tinnitus questions, 488 women reported experiencing daily persistent tinnitus lasting ≥5 minutes. Compared with women with no tinnitus (N=5989), women with persistent tinnitus were slightly older and were more likely to be postmenopausal, using oral postmenopausal hormone therapy, current or past smokers, and have type 2 diabetes, hypertension, and hearing loss.

**Table 1:** Nurses’ Health Study (NHS) and Nurses’ Health Study II (NHSII) participant characteristics according to tinnitus case-control status at the time of sample collection.

|  | **Controls**<br>**N=5989** | **Cases**<br>**N=488** |
| --- | --- | --- |
| **Age, years** | 51.76 (8.8) | 52.96 (7.9) |
| **Race, white, %** | 5666 (94.6) | 455 (93.2) |
| **Body mass index, kg/m<sup>2</sup>** | 25.52 (5.02) | 25.76 (4.52) |
| **Menopausal status, %** |  |  |
| Premenopausal | 2834 (47.3) | 198 (40.6) |
| Postmenopausal | 2968 (49.6) | 269 (55.1) |
| Indeterminate | 187 (3.1) | 21 (4.3) |
| **Current hormone use, %** |  |  |
| Not using any PMH | 2892 (48.3) | 196 (40.2) |
| Oral PMH | 1907 (31.8) | 199 (40.8) |
| Other PMH types | 33 (0.6) | 6 (1.2) |
| Premenopausal or missing | 1157 (19.3) | 87 (17.8) |
| **Current oral contraceptive use, %** |  |  |
| No | 5829 (97.3) | 481 (98.6) |
| Yes | 140 (2.3) | 7 (1.4) |
| missing | 20 (0.3) | 0 (0.0) |
| **Diabetes (ever), %** | 889 (14.9) | 86 (17.6) |
| **Hypertension (ever), %** | 2472 (41.3) | 222 (45.5) |
| **Depression, (ever) %** | 1737 (29.1) | 128 (26.2) |
| **Smoking, %** |  |  |
| Never | 3350 (56.1) | 262 (53.8) |
| Past | 2087 (35.0) | 179 (36.8) |
| Current | 534 (8.9) | 46 (9.4) |
| **Physical activity, MET-hours/week** | 17.32 (22.2) | 16.77 (19.9) |
| **DASH dietary score, %** |  |  |
| Quintile 1 | 1455 (24.3) | 101 (20.7) |
| Quintile 2 | 1196 (20.0) | 99 (20.3) |
| Quintile 3 | 1042 (17.4) | 99 (20.3) |
| Quintile 4 | 1180 (19.7) | 106 (21.7) |
| Quintile 5 | 1106 (18.5) | 83 (17.0) |
| **Alcohol intake, %** |  |  |
| None | 2214 (37.0) | 182 (37.3) |
| 1-4.9 g/d | 1945 (32.5) | 168 (34.4) |
| 5-14.9 g/d | 1045 (17.5) | 73 (15.0) |
| 15-29.9 g/d | 335 (5.6) | 29 (5.9) |
| 30+ g/d | 154 (2.6) | 17 (3.5) |
| Missing | 286 (4.8) | 19 (3.9) |
| **Cohort, NHSII, %** | 2178 (36.4) | 157 (32.2) |
| **Hearing loss (%)** |  |  |
| No | 3928 (65.6) | 129 (26.4) |
| Mild | 1378 (23.0) | 196 (40.2) |
| Moderate | 543 (9.1) | 122 (25.0) |
| Severe | 132 (2.2) | 38 (7.8) |
| Missing | 8 (0.1) | 3 (0.6) |

PMH: Postmenopausal hormone therapy
MET: Metabolic equivalent of task
DASH: Dietary Approaches to Hypertension; presented quintiles were calculated based on distribution in the entire cohort.
<sup>1</sup>Depression is defined as the use of anti-depressants in the past 2 years or self-reported diagnosis.

### Individual metabolites and metabolite classes associated with tinnitus

The analysis of 466 individual metabolites revealed several significant associations. In the fully -adjusted model (model 4), eight metabolites were associated with tinnitus at NEF-corrected p-value (NEF-p) <0.05 (**Figure 1 and Supplementary Table 2**). Homocitrulline [odds ratio (OR) and 95% confidence interval (CI) = 1.32 (1.16-1.50); NEF-p=0.01], C38:6 phosphatidylethanolamine [PE; OR (95%CI) = 1.24 (1.12-1.38); NEF-p=0.01], C52:6 triglyceride [TAG; OR (95%CI) = 1.22 (1.10-1.36); NEF-p=0.03], C36:4 PE [OR (95%CI) = 1.22 (1.10-1.35); NEF-p=0.03], C40:6 PE [OR (95%CI) = 1.22 (1.09-1.35); NEF-p=0.04], and C56:7 TAG [OR (95%CI) = 1.21 (1.09-1.34); NEF-p=0.04] were positively associated, whereas alpha-keto-beta-methylvalerate [OR (95%CI) = 0.68 (0.56-0.82); NEF-p=0.01] and levulinate [OR (95%CI) = 0.60 (0.46-0.79); NEF-p=0.04] were inversely associated with tinnitus. An additional 12 metabolites (6 TAGs, 4 PEs, 1 diglyceride (DAG), and thiamine) were positively associated, while an additional 2 lysophosphatidylcholines (LPC) were inversely associated with tinnitus at NEF-p<0.2. Odds ratios and significance levels were similar across the four logistic regression models we tested. Supplementary Figure 1 shows pairwise Spearman correlations across all metabolites associated with tinnitus at NEF<0.2. Similar associations were observed in sensitivity analyses in which we restricted to women who participated in the first NHS collection, the second NHS collection, the first NHSII collection, or to controls as defined within an individual study (i.e., controls in the breast cancer prospective case-control study; data not shown).

**Figure 1:**
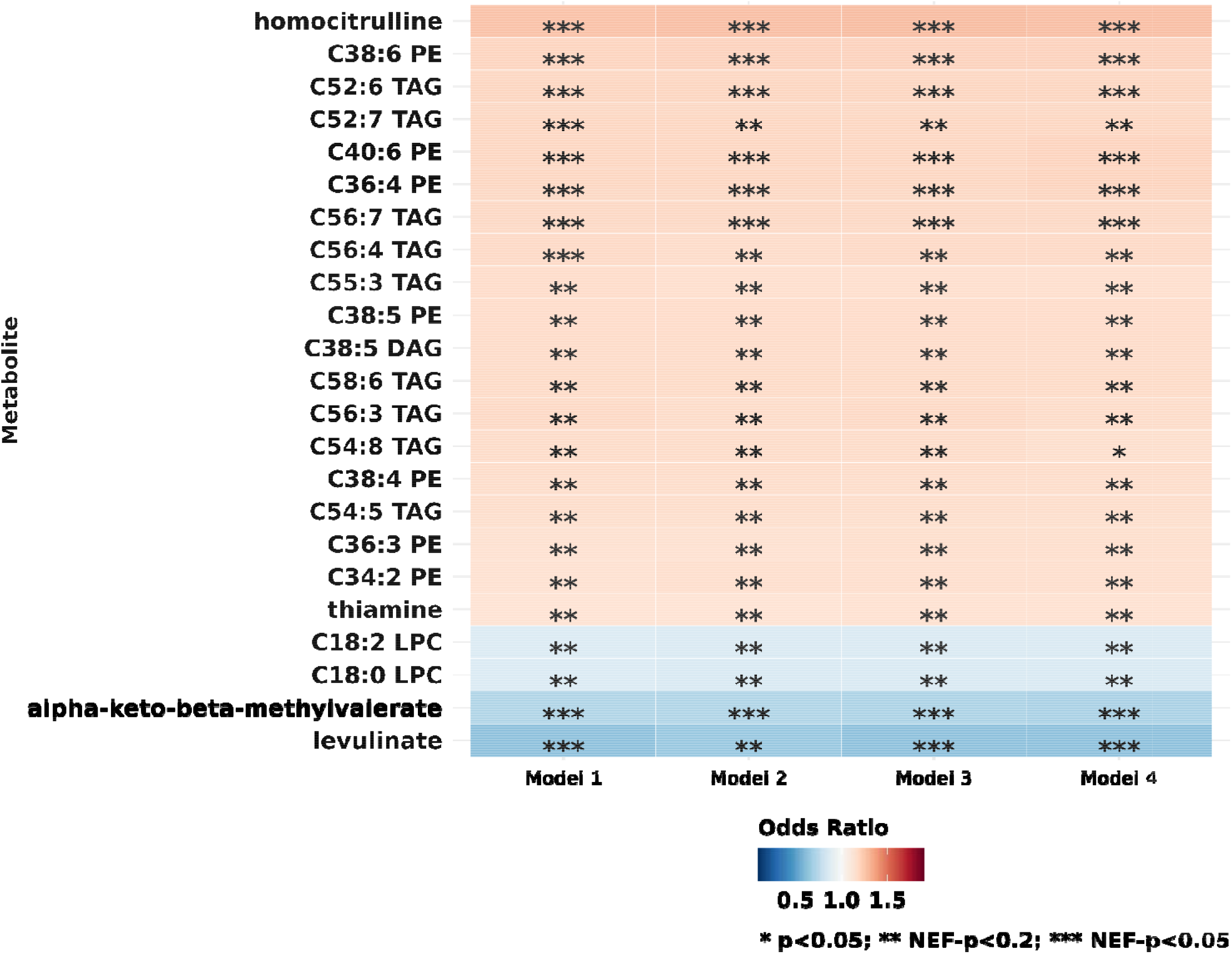
Individual metabolites associated with persistent daily tinnitus among 488 cases and 5989 controls in the Nurses’ Health Study and Nurses’ Health Study II at NEF-p<0.2 in at least one model. Positive associations are shown in shades of red while inverse associations are shown in shades of blue. Odds ratios of tinnitus were calculated per one standard deviation increase in probit transformed metabolite values. Statistical significance is overlaid on the plot: * Number of effective tests adjusted p value (NEF-p) <0.2; ** NEF-p <0.05. **Model 1** adjusts for age, race, fasting status, season of blood draw, collection, cohort, endpoint; **Model 2** adjusts for all covariates in Model 1+ body mass index; **Model 3** adjusts for all covariates in Model 2 + menopausal status + current oral hormone use, current oral contraceptive use; **Model 4** adjusts for all covariates in Model 3 + smoking status, physical activity, DASH dietary score, alcohol intake. PE: phosphatidylethanolamines; TAG: triglycerides; DAG: diglycerides; LPC: lysophosphatidylcholines; Caa:b: the fatty acyl chain(s) includes aa Carbon atoms and b double bonds.

In total, 337 of the 466 metabolites were mapped to 34 metabolite classes. Of the 34, 22 metabolite classes included at least 3 metabolites and were tested with MSEA. TAGs [normalized enrichment score (NES)=2.68; FDR<0.01], PEs (NES=2.48; FDR<0.01), and DAGs (NES=1.66; FDR=0.05) were positively associated while phosphatidylcholine (PC) plasmalogens (NES=-1.90; FDR=0.02), LPCs (NES=-2.23; FDR<0.01), and cholesteryl esters (CE; NES=-2.30; FDR<0.01) were inversely associated with tinnitus at FDR<0.05 (**Figure 2 and Supplementary Table 3**). PCs were positively associated, whereas alkaloids and derivatives, carboxylic acids and derivatives, phosphatidylethanolamine (PE) plasmalogens, carnitines, and lysophosphatidylethanolamines (LPE) were inversely associated with tinnitus at FDR<0.2.

**Figure 2:**
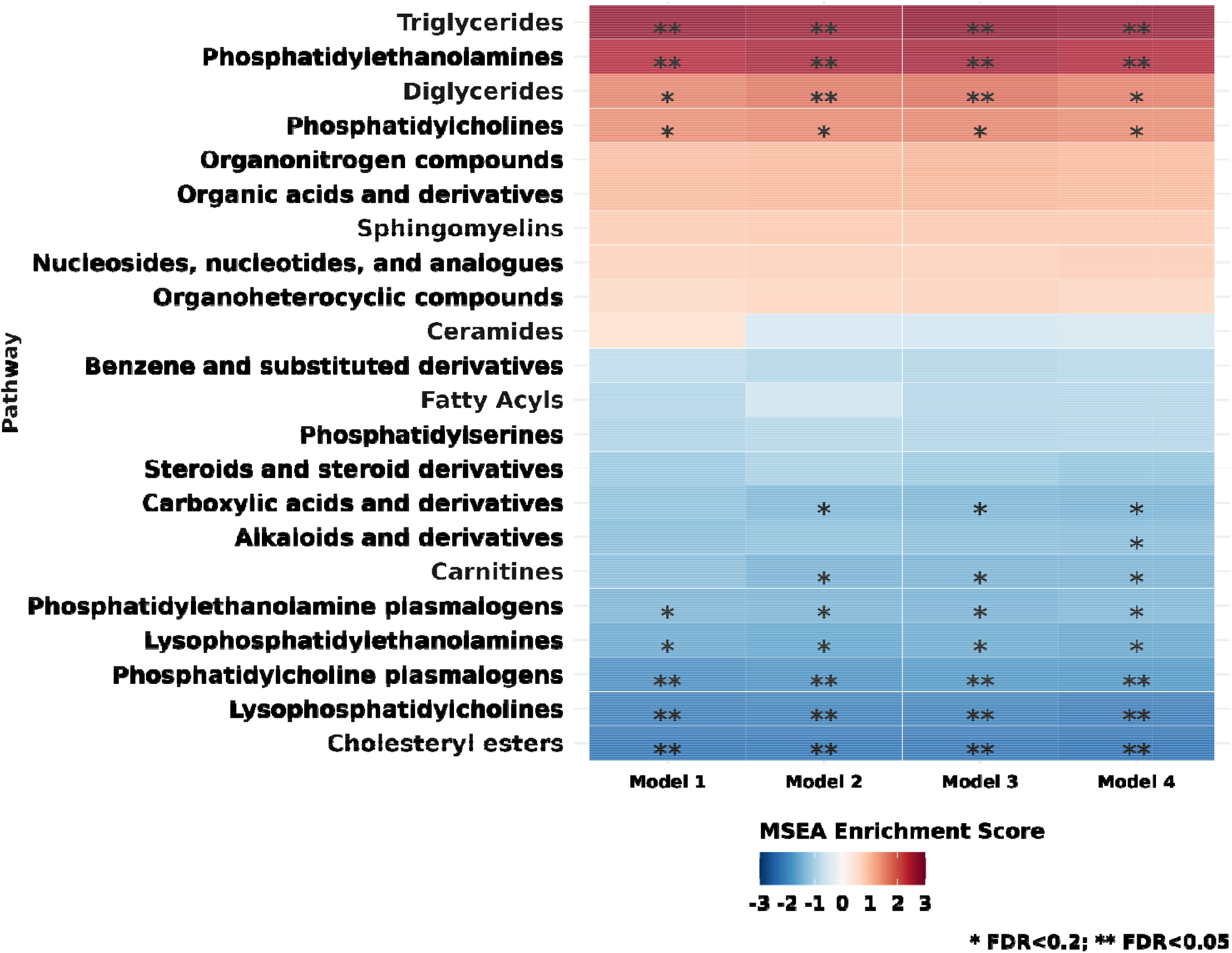
Metabolite classes associated with persistent daily tinnitus among 488 cases and 5989 controls in the Nurses’ Health Study and Nurses’ Health Study II. Positive associations are shown in shades of red while inverse associations are shown in shades of blue. Statistical significance is overlaid on the plot: * False discovery rate (FDR)<0.2; ** FDR<0.05. **Model 1** adjusts for age, race, fasting status, season of blood draw, collection, cohort, endpoint; **Model 2** adjusts for all covariates in Model 1+ body mass index; **Model 3** adjusts for all covariates in Model 2 + menopausal status + current oral hormone use, current contraceptive use; **Model 4** adjusts for all covariates in Model 3 + smoking status, physical activity, DASH dietary score, alcohol intake. MSEA: Metabolite Set Enrichment Analysis

### Individual metabolites and metabolite classes associated with tinnitus, according to hearing status

Among study participants, 129 cases and 3928 controls reported no hearing loss, while 160 cases and 675 controls reported moderate or severe hearing loss. Based on the fully multivariable-adjusted model 4, the observed associations with tinnitus among those with no hearing loss were statistically significant and stronger than among those with moderate or severe hearing loss for the following individual metabolite ORs: vitamin A [1.89 (1.29-2.77), NEF-p=0.19 vs. 0.84 (0.6-1.19), NEF-p=1], C52:6 TAG [1.46 (1.20-1.77), NEF-p=0.02 vs. 1.17 (0.96-1.43), NEF-p=1], C52:5 TAG [1.62 (1.30-2.02), NEF-p<0.01 vs. 1.06 (0.81-1.37), NEF-p=1], C36:3 DAG [1.41 (1.16-1.72), NEF-p=0.10 vs. 1.17 (0.95-1.44), NEF-p=1], C52:4 TAG [1.39 (1.14-1.69), NEF-p=0.19 vs. 1.18 (0.96-1.46), NEF-p=1], C20:4 CE [0.70 (0.57-0.85), NEF-p=0.07 vs. 0.87 (0.72-1.05), NEF-p=1], and C20:3 CE [0.69 (0.57-0.84), NEF-p=0.04 vs. 0.85 (0.71-1.03), NEF-p=1; **Figure 3 and Supplementary Table 4**]. In contrast, the association between alpha-keto-beta-methylvalerate and tinnitus was stronger and statistically significant among those with hearing loss [0.24 (0.10-0.54), NEF-p=0.11] than among those with no hearing loss [0.70 (0.50-0.98), NEF-p=1].

**Figure 3:**
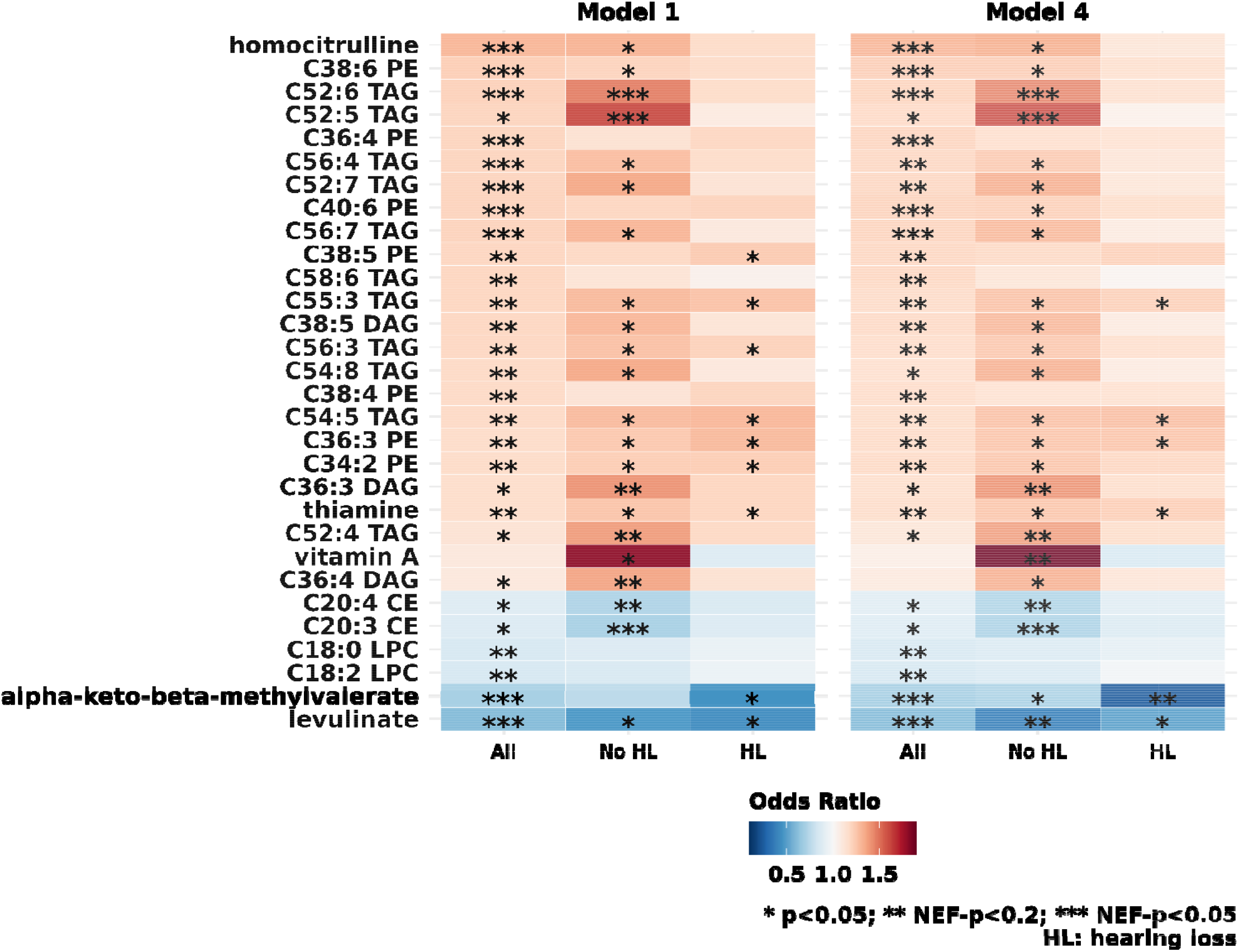
Individual metabolites associated with persistent daily tinnitus, without (129 cases, 3928 controls) and with moderate or severe hearing loss (160 cases, 675 controls), in the Nurses’ Health Study and Nurses’ Health Study II. This figure includes all metabolites identified in the main analysis and all metabolites with NEF-p<0.2 in the stratified analyses. The column **All** shows results from the main analysis which includes all tinnitus cases and controls. The column **No HL** shows results from the analysis restricted to those without hearing loss. The column **HL** shows results from the analysis restricted to those with moderate or severe hearing loss. Positive associations are shown in shades of red while inverse associations are shown in shades of blue. Odds ratios of tinnitus were calculated per one standard deviation increase in probit transformed metabolite values. Statistical significance is overlaid on the plot: * p<0.05; ** Number of effective tests adjusted p (NEF-p) <0.2; *** NEF-p <0.05. **Model 1** adjusts for age, race, fasting status, season of blood draw, collection, cohort, endpoint; **Model 4** adjusts for age, fasting status, season of blood draw, collection, cohort, endpoint, body mass index; menopausal status, current oral hormone use, current oral contraceptive use, smoking status, physical activity, DASH dietary score, alcohol intake. PE: phosphatidylethanolamines; TAG: triglycerides; DAG: diglycerides; CE: cholesteryl esters; LPC: lysophosphatidylcholines; Caa:b: the fatty acyl chain(s) includes aa Carbon atoms and b double bonds.

Based on the MSEA, TAGs and PEs were positively associated with tinnitus among those with and without hearing loss (**Figure 4 and Supplementary Table 5**). Based on the fully multivariable-adjusted model 4, the observed associations with tinnitus were stronger and statistically significant among those with no hearing loss than among those with moderate or severe hearing loss for the following metabolite classes: DAGs (NES=1.92, FDR=0.02 vs. NES=0.86, FDR=1), organic acids and derivates (NEF=1.47, FDR=0.16 vs. NES=-0.33, FDR=1), PC plasmalogens (NES=-1.80, FDR=0.04 vs. NES=-1.02, FDR=0.93), LPCs (NES=-1.96, FDR=0.01 vs. NES=-1.43, FDR=0.41), and CEs (NES=-2.49, FDR<0.01 vs. NES=-1.47, FDR=0.40). Steroids and steroid derivatives were associated with tinnitus among those with moderate or severe hearing loss (NES=1.86, FDR<0.01), but not among those with no hearing loss (NES=-1.47, FDR=0.28). Although LPCs and CEs were associated with tinnitus among those with moderate or severe hearing loss based on the least adjusted model (model 1), the associations were no longer statistically significant in the fully multivariable-adjusted model.

**Figure 4:**
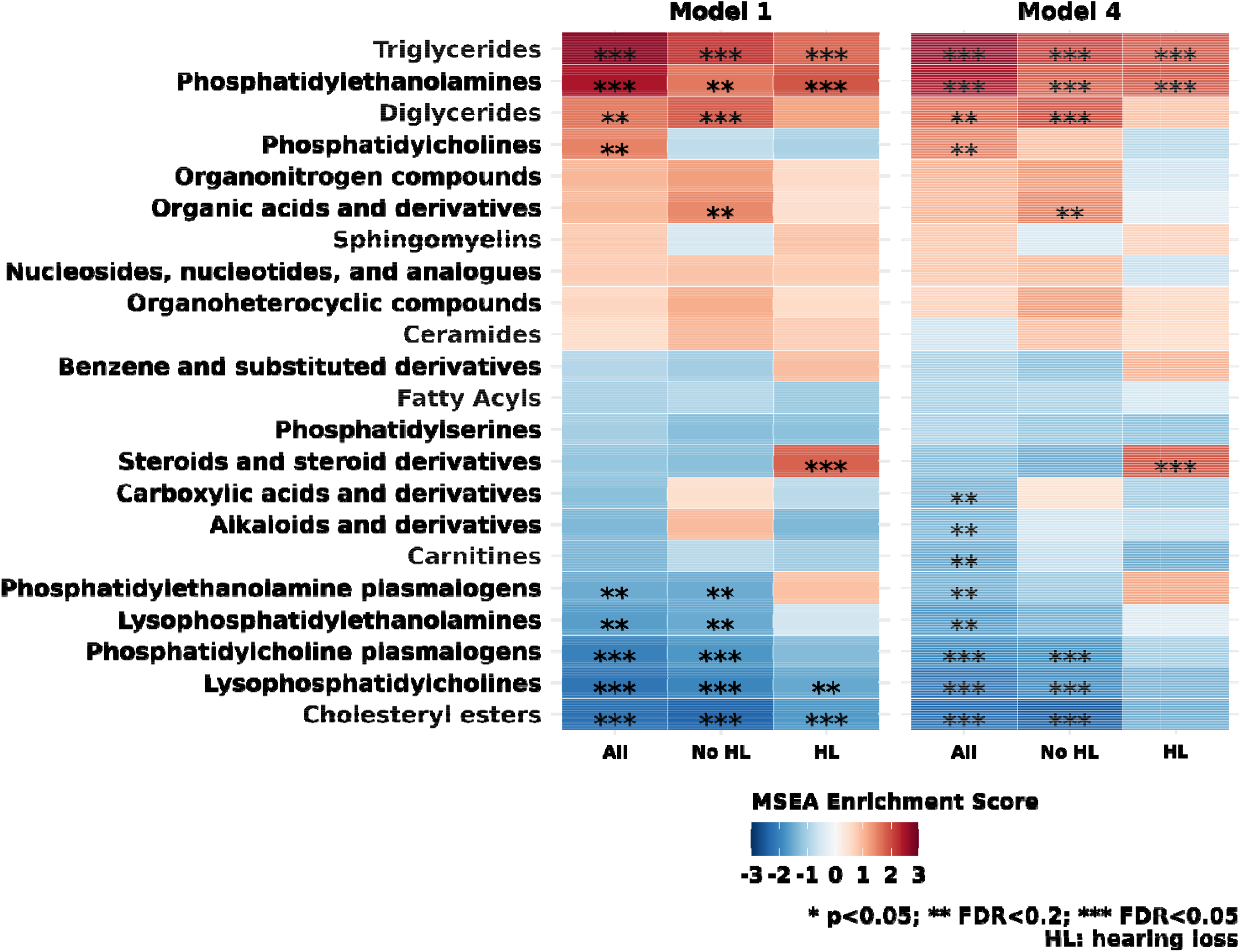
Metabolite classes associated with persistent daily tinnitus, without (129 cases, 3928 controls) and with moderate or severe hearing loss (160 cases, 675 controls), in the Nurses’ Health Study and Nurses’ Health Study II. NHS/NHSII. The column **All** shows results from the main analysis which includes all tinnitus cases and controls. The column **No HL** shows results from the analysis restricted to those without hearing loss. The column **HL** shows results from the analysis restricted to those with moderate or severe hearing loss. Positive associations are shown in shades of red while inverse associations are shown in shades of blue. Statistical significance is overlaid on the plot: * False discovery rate (FDR)<0.2; ** FDR<0.05; *** FDR<0.001. **Model 1** adjusts for age, race, fasting status, season of blood draw, collection, cohort, endpoint; **Model 4** adjusts for age, race, fasting status, season of blood draw, collection, cohort, endpoint, body mass index; menopausal status, current oral hormone use, current oral contraceptive use, smoking status, physical activity, DASH dietary score, alcohol intake. MSEA: Metabolite Set Enrichment Analysis

Due to the hypothesis generating nature of this study, all metabolites associated with tinnitus at p<0.05 based on the fully multivariable-adjusted model 4 are shown in **Figure 5** and among those without hearing loss in **Figure 6**.

**Figure 5:**
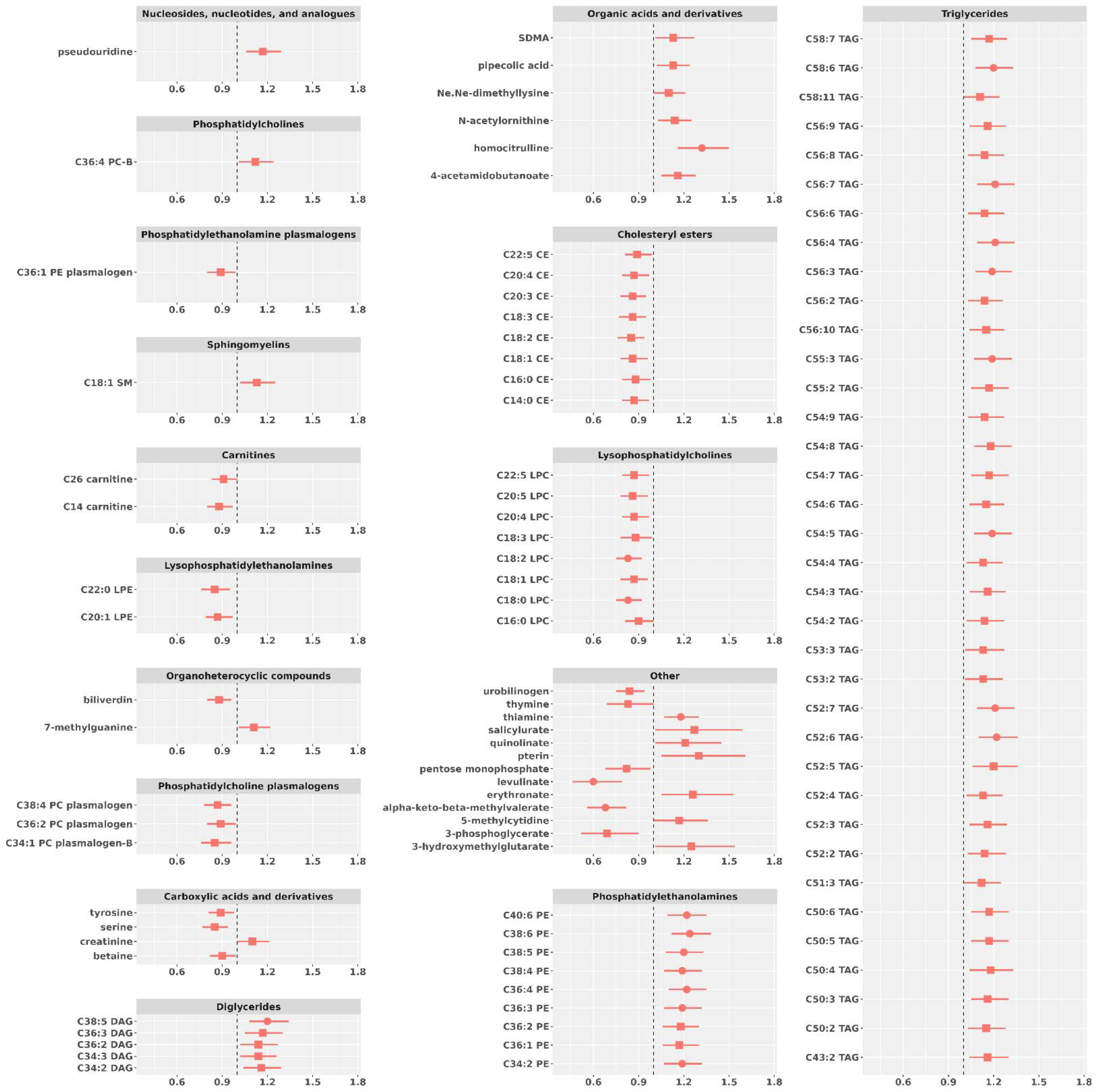
All individual metabolites associated with persistent daily tinnitus at p<0.05 among 488 cases and 5989 controls in the Nurses’ Health Study and Nurses’ Health Study II. Odds ratios of tinnitus were calculated per one standard deviation increase in probit transformed metabolite values. Metabolites with p-value<0.05 are shown as squares. Metabolites with NEF-p<0.2 are shown as circles. Model adjusts for age, race, fasting status, season of blood draw, collection, cohort, endpoint, body mass index; menopausal status, current oral hormone use, current oral contraceptive use, smoking status, physical activity, DASH dietary score, alcohol intake. PE: phosphatidylethanolamines; LPE: lysophosphatidylethanolamines; PC: phosphatidylcholines; LPC: lysophosphatidylcholines; SM: sphingomyelins; CE: cholesteryl esters; TAG: triglycerides; DAG: diglycerides; Caa:b: the fatty acyl chain(s) includes aa Carbon atoms and b double bonds.

**Figure 6:**
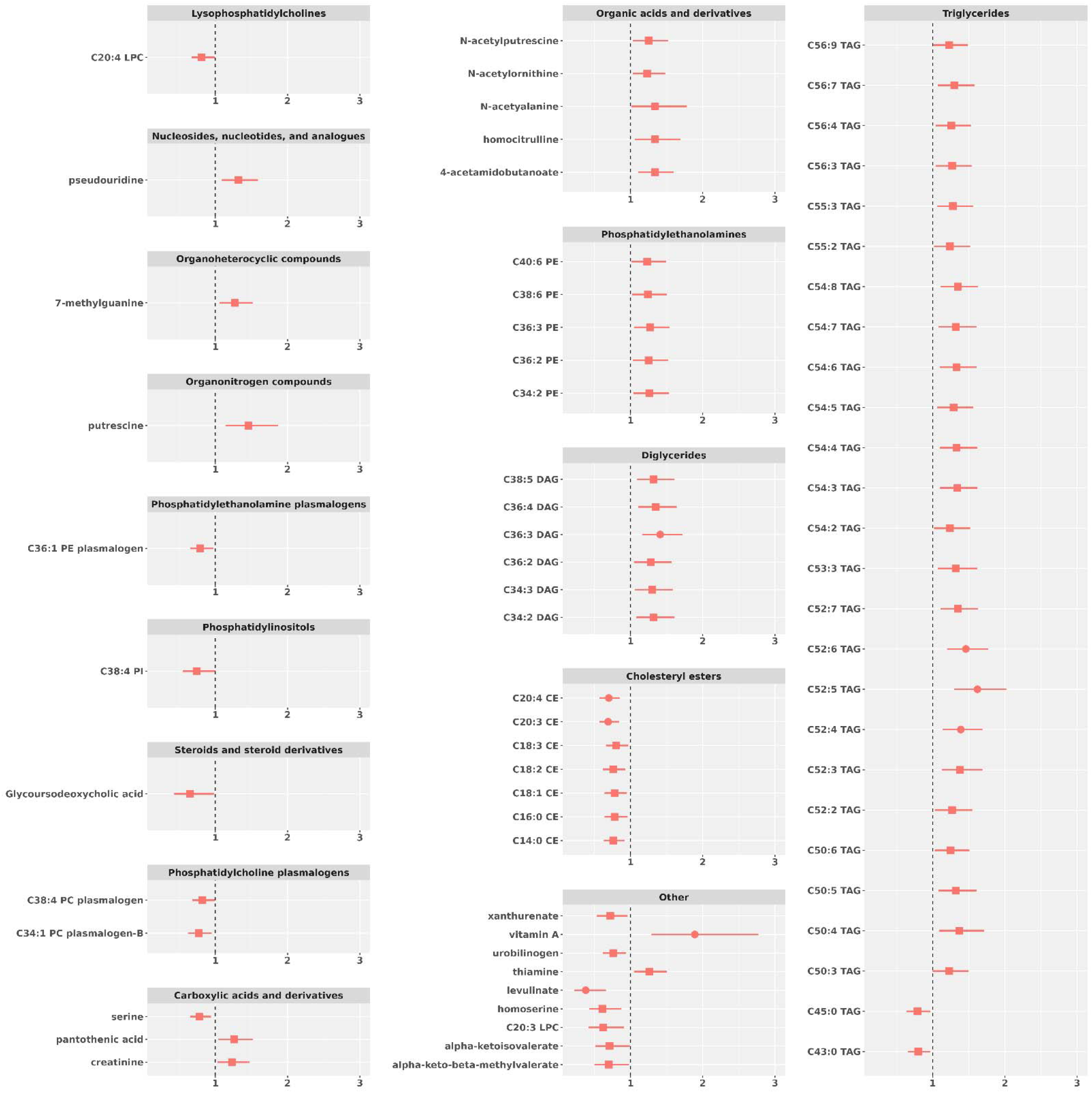
All individual metabolites associated with persistent daily tinnitus at p<0.05 among those without hearing loss (129 cases, 3928 controls) in the Nurses’ Health Study and Nurses’ Health Study II. Odds ratios of tinnitus were calculated per one standard deviation increase in probit transformed metabolite values. Metabolites with p-value<0.05 are shown as squares. Metabolites with NEF-p<0.2 are shown as circles. Model adjusts for age, race, fasting status, season of blood draw, collection, cohort, endpoint, body mass index; menopausal status, current oral hormone use, current oral contraceptive use, smoking status, physical activity, DASH dietary score, alcohol intake. PE: phosphatidylethanolamines; LPE: lysophosphatidylethanolamines; PC: phosphatidylcholines; LPC: lysophosphatidylcholines; SM: sphingomyelins; CE: cholesteryl esters; TAG: triglycerides; DAG: diglycerides; Caa:b: the fatty acyl chain(s) includes aa Carbon atoms and b double bonds.

## Discussion

In this large population-based metabolomic investigation of plasma metabolite profiles and tinnitus among women, we conducted a broad search for plasma biomarkers for tinnitus using a high-throughput, agnostic metabolomic approach. Overall, 466 plasma biomarkers were assessed in 6477 participants. We identified significant associations between a number of individual metabolites and metabolite classes and persistent tinnitus. Metabolites positively associated with persistent tinnitus included the amino acid homocitrulline, two three PEs (C38:6 PE, and C36:4 PE, C40:6), and two TAGs (C56:7 TAG C52:6 TAG), whereas alpha-keto-beta-methylvalerate and levulinate were inversely associated. The metabolites classes TAGs, PEs, and DAGs were positively associated, while PC plasmalogens and CEs were inversely associated with tinnitus. To our knowledge, this is the first large population-based plasma metabolomic study of tinnitus. Our findings suggest that metabolomic profiles could be a promising approach to identify tinnitus biomarkers and gain valuable insights into tinnitus pathophysiology.

We observed some striking differences in metabolite profiles among women who reported moderate or severe hearing loss compared with women with no hearing loss. For example, significant associations for certain metabolites and metabolite classes were observed among women with tinnitus and no hearing loss, but not among those with tinnitus and moderate or severe hearing loss. Specifically, several TAGs and vitamin A were positively associated with tinnitus, while CEs were inversely associated, but only among women with no hearing loss. For metabolite classes, positive associations with tinnitus were observed for diglycerides and organic acids, while inverse associations were observed for PC plasmalogens, LPCs and cholesteryl esters, but only among those with no hearing loss. In contrast, a strong positive association with tinnitus was observed for steroids and steroid derivatives only among those with moderate or severe hearing loss. Notably, strong positive associations for triglycerides and PEs and tinnitus were observed in both groups.

Persistent tinnitus is a common and often disabling condition ^27^ yet the complex interactions between individual-level and environmental factors that influence tinnitus generation and persistence are not fully understood. Tinnitus is a heterogenous disorder and distinguishing tinnitus phenotypes has been a clinical and research challenge. Metabolomic markers and pathways have been identified for cardiovascular outcomes ^28,29^ and several neurodegenerative conditions, including Alzheimer’s dementia (AD), Parkinson’s disease (PD) and multiple sclerosis (MS), ^30^ but the metabolomics of auditory disorders is still an emerging field. A metabolomic study in rats exposed to acoustic trauma sufficient to result in tinnitus and hyperacusis-related behaviors identified alterations in metabolic pathways involved in the urea, tricarboxylic acid and methionine cycles, and in purine and pyrimidine metabolism. Notably, several metabolite alterations demonstrated in brain tissue samples were also demonstrated in plasma. ^9^ However, metabolomic profiles and metabolic pathways can vary widely even among laboratory rodent strains commonly used in biomedical research, ^31^ and whether findings from animal models translate to humans is uncertain. ^32^ Nevertheless, the findings from animal studies demonstrate that there are significant metabolomic alterations associated with tinnitus and underscore the potential for human metabolomic studies to provide important insights into tinnitus pathophysiology.

Our findings indicate that metabolic dysregulation may contribute to the etiology of tinnitus, although further research is needed to clarify which metabolites and metabolic pathways may be most influential. Tinnitus is a complex disorder and metabolic perturbations related to hyperlipidemia, atherosclerosis, hypertension, diabetes, genetic and environmental factors, and how these relate to inflammation, oxidative stress, neural transmission and neuroplasticity, may all play a role. We identified significant associations with several metabolites and metabolite classes that suggest dysregulation of lipid metabolism and other metabolic pathways may be influential; the significant findings and potential mechanisms are briefly discussed below. Our study substantially augments the currently scarce literature on plasma metabolomic biomarkers for tinnitus. Specifically, a few small metabolomic investigations of sensorineural hearing loss in animals and humans have examined metabolomic profiles in perilymph, plasma, urine, temporal bone or brain tissue, ^33^ and evaluated metabolomic alterations following noise or cisplatin exposure. Plasma metabolomic alterations following noise exposure were observed for several individual metabolites and pathways, such as those involving glycerophospholipid, choline and fatty acid metabolism.^34^

### Homocitrulline

Homocitrulline, which was positively associated with tinnitus in our study, is an amino acid and a by-product of ornithine metabolism. Homocitrulline is formed in mitochondria from lysine and carbamoyl phosphate; plasma concentration reflects the overall carbamylation rate of serum proteins. Brain degenerative protein modifications, such as carbamylation, have been associated with neuroinflammation and proteinopathy. ^35^ Carbamylation irreversibly neutralizes the positively charged lysine residue ^36^ and can lead to altered function and pathogenic conformation. ^37,38^ Along with leading to protein denaturation and proteolysis, homocitrulline may be recognized by the immune system as a non-naturally coded amino acid ^39,40^ and promote autoimmune and inflammatory responses. ^35,41^ Plasma homocitrulline levels are elevated in individuals with ornithine translocase (SLC25A15) deficiency, which is associated with neurologic abnormalities including developmental delay, ataxia, spasticity, learning disabilities, cognitive deficits and seizures. ^42^ Altered plasma homocitrulline levels were identified among individuals with utism spectrum disorder. ^43^ In addition, higher plasma homocitrulline was associated with chronic kidney disease, ^44^ liver disease, ^45,46^ coronary artery disease, ^47^ and higher risk of mortality among Type II diabetics. ^48^ Homocitrulline may be involved in neuroinflammation; evidence suggests homocitrulline triggers early activation of proinflammatory cells, autoimmune inflammation ^49^ and progression of neuroinflammation. ^50^ Homocitrulline also promotes endothelial dysfunction and vascular disease, ^51^ possibly indicating a neurovascular component of tinnitus pathophysiology.

### Triglycerides (TAGs) and diglycerides (DAGs)

Lipids play a fundamental role in the pathophysiology of neurodegenerative diseases. ^52,53^ Specific lipid species are structural components of cellular membranes (e.g., cholesterol and sphingolipids) and regulate a range of critical aspects of brain function. ^54^ A number of TAGs and DAGs, key components of fatty acid metabolism, were positively associated with tinnitus in our study. These findings are consistent with a small case-control study in Turkey that compared serum lipid values among 91 individuals with tinnitus with 65 age-and sex-matched controls without tinnitus and found higher serum TAG, total cholesterol, and LDL among those with tinnitus. ^55^ An intriguing observation in our study was the strong significant association of higher plasma TAGs with tinnitus among individuals with and without hearing loss, while higher plasma DAGs were significantly associated with tinnitus only among those without hearing loss. Evidence suggests that perturbations in lipid metabolism are associated with several neurodegenerative disorders, including amyotrophic lateral sclerosis (ALS), ^56-58^ PD, ^52,59^ and AD. ^53,60^ Whether lipidomic profiles may aid in the distinction of tinnitus sub-groups and whether targeted interventions that modify plasma lipid status may influence the risk and course of tinnitus merit further exploration.

### Phosphatidylethanolamine (PE)

PEs, a class of phospholipids found in biological membranes, are synthesized by the addition of cytidine diphosphate-ethanolamine to diglycerides, releasing cytidine monophosphate. The amine of phosphatidylethanolamines can subsequently be methylated by S-Adenosyl methionine to yield phosphatidylcholines, found mainly in the inner (cytoplasmic) leaflet of the lipid bilayer. In our study, PEs were positively associated with tinnitus among those with and without hearing loss. PEs are the second-most abundant phospholipids in the cell and are involved in protein biogenesis, oxidative phosphorylation, membrane fusion, mitochondrial stability, and serve as precursors to other lipids. There are four independent PE biosynthesis pathways, three of which take place in the endoplasmic reticulum and one, the phosphatidylserine decarboxylase pathway, which takes place in the mitochondrion. ^61^ PE dysregulation has been implicated in neurodegenerative disease. ^53,62^ A small study among adults with AD found an inverse association of PEs with self-reported hearing loss, but the association was not significant after correction for multiple comparisons. ^62^ A study among men with occupational noise exposure (n=62 men with noise-induced hearing loss, n=62 controls) found lower plasma PE among those with hearing loss. ^63,64^

### Phosphatidylcholine (PC) plasmalogens

PC plasmalogens are common glycerophospholipids that rigidify membranes, facilitate signaling processes, and can act as free radical scavengers and protect membrane lipids from oxidation. ^65^ PC plasmalogens are formed via headgroup transfer from PE plasmalogens. Plasmalogens increase early in life, rapidly decrease with aging, and may play a role in neurodegenerative disease. ^66^ Alterations in plasma PC plasmalogens were demonstrated for several neurodegenerative conditions, including AD, PD and MS. ^66^ Plasma PC plasmalogens were also lower in individuals with ALS. ^67^ Increased lipid oxidation occurs in several inflammatory and neurodegenerative conditions and is associated with lower plasmalogen levels. Potential mechanisms for a neuroprotective influence of plasmalogens in these disorders include the facilitation of lipid signaling and protection of membrane lipids from oxidation. ^65^ A study among individuals with AD found significantly lower plasma PC among those with AD with hearing loss compared with those with AD without hearing loss. ^68^ The role of PC plasmalogens in tinnitus is unclear, but studies of age-related hearing loss in rats suggest that PC may play a protective role in cochlear mitochondrial function. ^69^ In our study, plasma PC plasmalogens were also inversely associated with tinnitus, thus further investigation of a potential protective role for PC plasmalogens in tinnitus prevention and management could be promising.

### Lysophosphatidylcholines (LPC)

LPCs are a class of lipids derived from the cleavage of PC by phospholipase A_2_ (PLA_2_) or by the transfer of fatty acids to free cholesterol via lecithin-cholesterol acyltransferase (LCAT) and are components of oxidatively damaged low-density lipoprotein. ^70-72^ The enzymatic cascade involved in LPC metabolism is complex, and the mechanisms underlying the physiological effects of LPCs are not fully understood. Through G protein-coupled receptor signaling, LPCs can promote inflammation, disrupt mitochondrial integrity, and induce apoptosis. ^73^ LPCs may adversely influence endothelial and vascular function and lead to progression of atherosclerosis and cardiovascular disease. However, LPCs may also help reduce plasma glucose. ^74,75^ Notably, in clinical studies, plasma LPCs were inversely associated with cardiovascular disease. ^76-78^ Plasma LPCs were also lower among individuals with AD, ^79,80^ and the LPC to PC ratio was lower in plasma and cerebrospinal fluid among those with AD. ^81,82^ Further, the ratio of plasma LPC to PC was found to distinguish AD from mild cognitive impairment. ^82^ It has been suggested that LPCs may improve cognition and have other neuroprotective effects, possibly by increasing docosahexaenoic acid (DHA), influencing signaling pathways that protect against apoptosis and enhancing production of nerve growth factor. ^83-87^ In our study, we observed an inverse association of plasma LPCs and persistent tinnitus, a novel finding consistent with previous studies suggesting that LPCs may help reduce inflammation and act as mediators in neuronal pathways, in addition to their role as glycerophospholipid metabolism intermediates. ^88-90^

### Cholesteryl esters (CEs)

We observed an inverse association of CEs and persistent tinnitus. Altered cholesterol metabolism and lower plasma CEs have been demonstrated in individuals with several neurodegenerative conditions, including PD, ^91^ AD, ^92,93^ ALS ^94^ and MS ^95,96^. Cholesterol serves as an essential membrane component, signaling molecular cofactor, and steroid precursor, and CEs are a major component of high density lipids (HDL). CEs are mostly synthesized in plasma by the transfer of fatty acids to cholesterol from PC, catalyzed by the enzyme LCAT (lecithin: cholesterol acyl transferase). Cholesterol esterification allows more cholesterol to be packaged into the interior of lipoproteins and increases the capacity and efficiency of cholesterol transport. In our study, plasma CEs were inversely associated with tinnitus among those with and without hearing loss, although the association was not statistically significant among those with hearing loss in the fully adjusted model.

Taken together, these findings indicating alterations in plasma TAGs, PCs, LPCs, PEs, and cholesterol-related metabolites suggest a role of lipid dysregulation in tinnitus pathogenesis and are consistent with findings for several other neurodegenerative disorders.

### Alpha-keto-beta-methylvalerate and levulinate

We observed inverse associations for two novel plasma metabolites not previously examined in relation to tinnitus, alpha-keto-beta-methylvalerate and levulinate. Alpha-keto-beta-methylvalerate is a catabolic product of iso-leucine, a branched-chain amino acid. It is used as a B-complex vitamin marker in lab tests, because B vitamins are necessary for the enzymatic catabolism of isoleucine ^97^. Levulinate is a food additive and is used as a calcium supplement source. Further studies to investigate the role of these two metabolites in the pathogenesis of tinnitus are needed.

### Strengths and Limitations

Strengths of our study include the use of two richly characterized cohorts, enabling adjustment for a broad range of covariates along with the use of a well-characterized metabolomics platform that measured a large set of metabolites with robust CVs and low missingness. Our study was limited by the use of non-random samples from previous NHS and NHS II metabolomic studies, the coverage of a subset of the metabolome by the used metabolomic profiling methods, potential unmeasured confounding, the cross-sectional nature of our study, and the use of one plasma sample to represent long term exposure. Previous examinations of the stability of metabolite profiles in NHS showed 90% of measured metabolites demonstrated reasonable within-person reproducibility over 1-2 years and over 10 years (most metabolites had a Spearman or intra-class correlation coefficient > 0.4 over 1-2 and over 10 years) showing that one plasma sample is able to represent long term exposure. ^21,98^ Due to the nature of the condition, subjective tinnitus is perceived only by the individual, therefore the diagnosis of tinnitus must rely on self-report. ^99^ Notably, among those who reported persistent tinnitus in 2009, 88% also reported persistent tinnitus again 4 years later on the 2013 questionnaire. Definitions of tinnitus used in previous studies have varied greatly. ^100^ We examined a stringent definition of tinnitus, daily tinnitus lasting ≥ 5 minutes, to reduce potential misclassification and to evaluate cases of tinnitus that were most likely to be clinically meaningful. The study population included predominantly white female health care professionals, thus research in racially and ethnically diverse populations is warranted.

## Conclusion

In this first large population-based metabolomic study of tinnitus, we identified several plasma metabolites and metabolite classes that were significantly associated with persistent tinnitus, and the findings differed by hearing status. These findings provide new insights into metabolic pathways that may be involved in tinnitus etiology and suggest that metabolomic profiling offers a promising approach to identify tinnitus biomarkers and inform investigations of novel therapeutic targets for this challenging disorder.

## Data Availability

The demographics and questionnaire data together with the plasma metabolomics profiles are not publicly available for the following reason: data contain information that could compromise research participant privacy. Reasonable requests to access these data should be made via http://www.nurseshealthstudy.org/researchers.

## Funding information

This research was funded by the Royal National Institute for Deaf People (RNID) Grant G103_Curhan, Massachusetts Eye and Ear Foundation, Allan and Muriel Greenblatt Philanthropic Fund, Bertarelli Foundation Professorship and grants U01 DC010811, UM1 CA186107, P01 CA87969, R01 CA49449, R01 HL034594, R01 HL088521, R01 CA67262 and UO1 CA176726 from the National Institutes of Health. DBW reports being consult and/or on the Scientific Advisory Boards of Recursion Therapeutics, Inc, Science 24/7, NF2 BioSolutions, NF Bio, Daybreak Vision Project, Mulberry Therapeutics, Inc., Skylark Biotherapy, Inc., and Salubritus Bio, Inc. GCC reports being an employee of OM1, Inc, holding stock of Allena Pharmaceuticals, royalties (Section Editor and author) from UpToDate, and grant funding from Decibel Therapeutics.

## Notes

**Conflict of interest:** The authors declare that they have no conflict of interest.

### Author Declarations

The study protocol was approved by the Institutional Review Boards of the Brigham and Women's Hospital and Harvard T.H. Chan School of Public Health.

